# Changes in mobility patterns during the COVID-19 pandemic in Zambia: implications for the effectiveness of NPIs in Sub-Saharan Africa

**DOI:** 10.1101/2022.07.20.22277849

**Authors:** Stacie Loisate, Simon Mutembo, Rohan Arambepola, Kabondo Makungo, Elliot N Kabalo, Nyambe B. Sinyange, Nathan Kapata, Mazyanga Liwewe, Andrew Silumezi, Gershom Chongwe, Natalya Kostandova, Shaun Truelove, Amy Wesolowski

## Abstract

The COVID-19 pandemic has impacted many facets of human behavior, including human mobility partially driven by the implementation of non-pharmaceutical interventions (NPIs) such as stay at home orders, travel restrictions, and workplace and school closures. Given the importance of human mobility in the transmission of SARS-CoV-2, there have been an increase in analyses of mobility data to understand the COVID-19 pandemic to date. However, despite an abundance of these analyses, few have focused on Sub-Saharan Africa (SSA). Here, we use mobile phone calling data to provide a spatially refined analysis of sub-national human mobility patterns during the COVID-19 pandemic from March 2020-July 2021 in Zambia. Overall, among highly trafficked intra-province routes, mobility decreased up to 52% from March-May 2020 compared to baseline, which was also the time period of the strictest NPIs. However, despite dips in mobility during the first wave of COVID-19 cases, mobility returned to baseline levels and did not drop again suggesting COVID-19 cases did not influence mobility in subsequent waves.

## Introduction

The COVID-19 pandemic has resulted in an unprecedented global public health crisis that has drastically changed human behavior, particularly the travel patterns of individuals. At the start of the pandemic in early 2020, many non-pharmaceutical interventions (NPIs) were put in place to minimize the spread of SARS-CoV-2. [1] Some NPIs including travel restrictions, business/school closures, and social distancing were implemented to varying degrees globally and have been shown to minimize the likelihood of onward transmission. [2] Spurred by the public health relevancy and importance of predicting the likely trajectory of SARS-CoV-2, datasets quantifying human mobility have also become more readily available. [3] This has led to a deluge of analyses focused on quantifying reductions in human mobility and implications for evaluating the effectiveness of NPIs. [4] However, despite global efforts, to date there have been few analyses focused on Sub-Saharan Africa (SSA). The pandemic has shown substantially different patterns in SSA compared to other regions with overall lower reported cases and mortality rates despite serological results suggesting overall high rates of infection. [5] Poor surveillance may be a root cause for the differences in the pandemic to date, however other factors such as protection from endemic coronaviruses, an overall younger average age resulting in less severe infections, and the early implementation of NPIs relative to the number of reported cases have been posited as possible reasons for some of the observed differences. [6] Disentangling these factors requires detailed data on human mobility and behavioral patterns from countries in SSA.

Understanding how human behavior has changed during NPIs in SSA has been explored, but primarily via aggregated data as a proxy for mobility such as Google Mobility Reports. [7] Given biases in who is represented in these data based on Google app usage, these data may be presenting a biased view of behavior. Further, these data often have spatial data gaps or are aggregated to represent mobility throughout the entire country, which may hinder detailed analyses highlighting sub-national patterns of transmission. In addition, spatial heterogeneity in transmission is further impacted by limited testing availability which limits approaches to understand transmission dynamics directly from confirmed case or death data. In Africa, vaccination remains limited with just under 16% of persons fully vaccinated, compared to some high-income settings where fully vaccinated proportions are greater than 60%. [8]–[10] In these settings, travel and social distancing restrictions may be more readily implemented to further stem the spread of SARS-CoV-2. Additionally, changes in the virus and the emergence of new variants alter our landscape of protection and susceptibility requiring NPIs to be implemented despite widespread vaccination. [11]

Using detailed data on mobility patterns quantified from mobile phone calling data, we provide the first spatially resolved analysis of mobility during the COVID-19 pandemic in Zambia from 01 March 2020 to 07 July 2021. We couple these data with information on reported cases, the implementation of various NPIs, and a model of mobility to understand where, how, and when travel has changed sub-nationally since March 2020. We further investigate implications for the effectiveness of travel restrictions moving forward as the course of the pandemic continues to spread through populations and prolong morbidity and mortality.

## Materials and Methods

### Data on mobility

Deidentified, anonymized, call detail records (CDRs) were obtained with permission from a leading mobile phone operator in Zambia. CDRs from 108 districts across all 10 Zambia provinces were included in the final data (Fig 1A). Individual subscribers were assigned a most used mobile phone tower per day based on the routing tower where most of their records were recorded. Trips were defined based on the most used mobile phone tower on subsequent days where a trip was counted if these two towers were different. If these two most used towers were the same, then this was recorded as a “stay” where a subscriber did not change their location. CDR data was processed further by aggregating individual movements both spatially by district and temporally by day. Aggregation was conducted at the district and province level to construct origin-destination matrices of movement between districts and provinces. Overall, between 566,685 – 871,851 unique mobile phone subscriber IDs were recorded per day from 1 March 2020 to 7 July 2021. Google Community Mobility Reports were obtained via open access from Google at https://www.google.com/covid19/mobility/. [12] The output from Google’s data focuses on national percent changes from baseline and is aggregated into several categories including changes in visits retail stores, grocery stores, parks, transit, workplace, as well as residential mobility. To compare with the mobile phone data, we only looked at the changes in workplace mobility as a proxy for overall national mobility.

**Fig 1.**
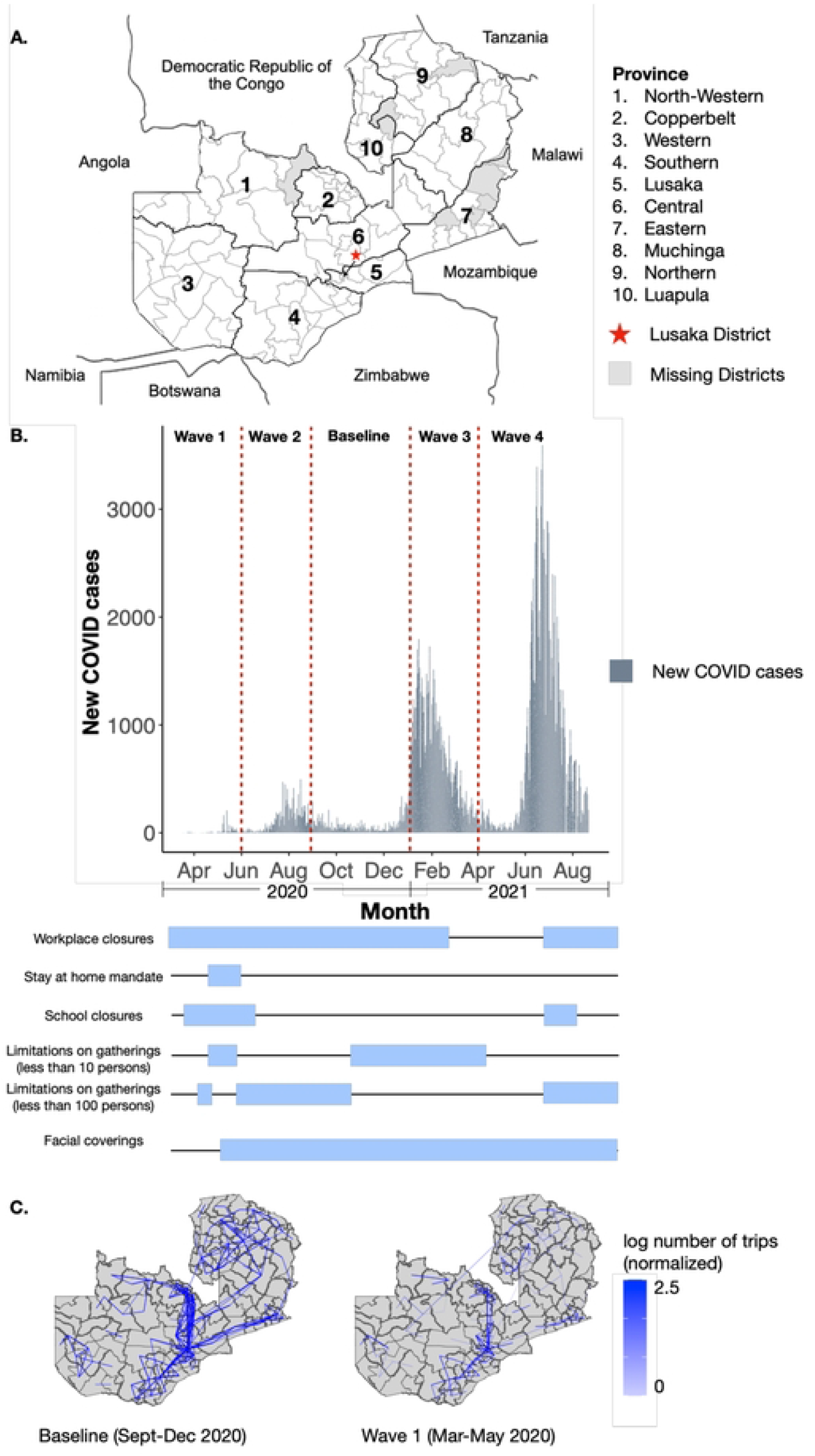
Overview of districts included, COVID-19 case counts, and baseline mobility. **(A)** The map of Zambia and bordering countries. Zambia Districts and Provinces (outlined in black) are labeled with districts without mobile phone data shaded in grey. **(B)** The time series of COVID-19 cases in Zambia from March 2020-August 2021 with the dates when key NPIs instated shown in blue bars. **(C)** The districts of Zambia displaying the log number of trips between districts for the top 10% most frequently traveled routes. Data were normalized to a fixed range between 0 and 1 by scaling to minimum and maximum values, and are shown for both baseline (September-December 2020) and Wave 1 (March-May 2020).

### COVID-19 data

Daily case data for Zambia was obtained from the Johns Hopkins University COVID-19 dashboard. Case data was available by district from 18 March 2020 to 22 February 2021 (Fig 1B). [13]

### Non-pharmaceutical interventions (NPIs)

Data on dates and duration of NPIs put into place during 2020 were obtained from the Oxford Covid-19 Government Response Tracker. [1] Initial NPIs were implemented nationally in Zambia from 20 March 2020 – 31 May 2020. The main restrictions included workplace and school closures, stay at home orders, and limitations on gatherings. The stringency level of NPIs in Zambia were the strictest between March and May 2020, but later varied throughout the course of the pandemic. NPIs implemented from 20 March – 31 December 2020 can be defined as:

| National Restrictions (<u>general</u> recommendations/restrictions) |  |  |  |
| --- | --- | --- | --- |
| Restriction | Definition | First Implementation | Second Implementation |
| School Closures | Required closing *all* levels | 20 Mar – 31 May 2020 |  |
|  | Required closing *some* levels | 01 Jun – 13 Sept 2020 | 14 Dec – 31 Dec 2020 |
|  | Recommended closing | 14 Sept – 13 Dec 2020 |  |
| Workplace Closures | Required closures for *all* workplaces except for essential businesses | 02 May – 07 May 2020 |  |
|  | Required closing and/or work from home for *some* businesses | 08 May – 19 Aug 2020 | 01 Sept – 31 Dec 2020 |
| Stay at Home Orders | Strong recommendation to stay home | 09 Apr – 14 Apr 2020 | 26 Sept – 31 Dec 2020 |
| Limitations on Gatherings | Limit gatherings to less than 100 persons | 26 Mar – 14 Apr 2020 | 08 May – 25 Sept 2020 |
|  | Limit gatherings to less than 10 persons | 26 Sept – 31 Dec 2020 |  |

| Sub-National Restrictions (<u>targeted</u> recommendations/restrictions) |  |  |  |
| --- | --- | --- | --- |
| Restriction | Definition | First Implementation | Second Implementation |
| Stay at Home Orders | Mandated orders to stay at home except for essential trips – Kafue District | 15 Apr – 07 May 2020 |  |
| Limitations on Gatherings | Limit gatherings to less than 10 persons – Kafue District | 15 Apr – 07 May 2020 |  |

### Analysis

The analysis in this manuscript largely focuses on describing human mobility in Zambia during the start of the COVID-19 pandemic. The primary areas of exploration are (1) describing overall changes in mobility, (2) comparing inter- and intra-province travel, (3) exploring trends between mobility and COVID-19 case data, and (4) comparing mobile phone data with Google Mobility Reports. We temporally aggregated the mobility data to align with the COVID-19 waves as follows: Wave 1: 01 March – 31 May 2020, Wave 2: 01 June – 31 Aug 2020, and Wave 3: 01 Jan – 31 Mar 2021. For some analyses, each wave was then averaged to obtain a district-level mean trips during that time period. Since 2019 mobile phone data was not available, the time period from 01 Sept – 31 Dec 2020, where mobility was stable and no NPIs were in place, was used as a proxy for “baseline” mobility. Since the mobile phone data did not show any drops in mobility during Waves 2 or 3, we focused our analysis on mobility and NPIs implemented during Wave 1. For analyses comparing mobility during the different waves and baseline, the data were normalized to the baseline values. For some analyses, Welch’s two-sample t-tests were used to compare groups.

### Mobility Model

To investigate the rate of decreasing mobility in the initial stage of the pandemic and the effect of explanatory variables on this decrease, we fit a Bayesian model of mobility to the observed trip counts. For each pair of districts, the rate of trips from district *i* to district *j* on day *t, λ*_*ijt*_, was modelled as a fraction of the baseline travel rate for this pair, *r*_*ij*_,

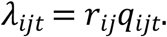

The fraction *q*_*ijt*_ represented the cumulative daily decrease in travel in the district, modified by the effect of explanatory variables,

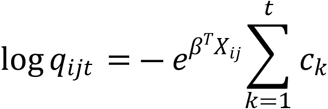

where *c*_*k*_ > 0 was the decrease in travel on day *k, X*_*ij*_ were explanatory variables and *β* was a vector of effects to be learned. Euclidean distance was used as the primary explanatory variable in this analysis. A Poisson likelihood was used for the number of trips observed each day from district *i* to district *j*,

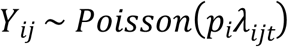

where *p*_*i*_ was the population in district *i* and the model was completed by applying priors to the model parameters. Rates of decrease were parameterized by on the log scale with an AR(1) prior, ϕ = 1, c = 0, and ε = 0.5.The model was implemented in the R programming language using the TMB package and maximum a posteriori estimates were obtained using the Laplace approximation. [14]

### Transmission Model

To compare the impacts of different mobility information on simulations of SARS-CoV-2 transmission dynamics, we used a spatial semi-mechanistic time-series transmission model that integrated each mobility data set. [15] Key parameters in the model were M, α, and β_0_; where M represents the mobility matrix to approximates connectivity between districts, α represents a scaling parameter to account for the discrete time steps, and the β_0_ represents the transmission coefficient, which can be approximated by R_0_, which we are assuming to be 2 for this analysis. [16] For our model we will set α to 0.99 suggesting near homogenous mixing patterns has been done for other similar models. [15], [17] We investigated the impact on SARS-CoV-2 cases based on two mobility matrices informed by either the Google Mobility Reports or the mobile phone data. Mobility matrices were generated by (1) determining the national average reduction of mobility in Zambia during Wave 1, (2) reducing baseline mobility by the reduction factor determined in the previous step, and (3) normalizing the data. The transmission model required the mobility matrix to be a fixed range between 0 and 1 by so the data was scaled using the minimum-maximum scaling methods.

The number of persons susceptible (*S*_*i,t*_) or infected (*I*_*i,t*_) at each time point, *i* (one time point = one week), for a district, *j*, can be estimated by:

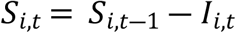

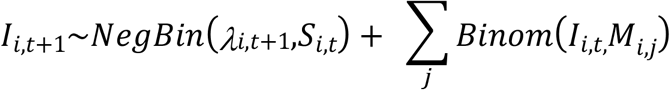

Where,

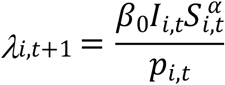

Each model was run fifty times and averaged for further analysis. We also estimated the time until the first 10, 50, and 100 cases were reached in each district and compared output between mobility datasets and stratified by traffic density.

### Ethics statement

Study approval was granted by the Johns Hopkins Bloomberg School of Public Health (IRB 15892) which deemed the work exempt (and hence consent was not obtained).

## Results

### Mobility decreased nationally following the implementation of NPIs

At baseline (averaged from 01 September – 31 December 2020) a total of 880,337 average daily trips were recorded between districts (range 1-200,277 daily trips per route) by the mobile phone data. Most trips (46%) occurred within a province with trips from the capital province, Lusaka, accounting for 12% of total trips. As in other settings, travel greatly decreased (40.8%, IQR: 21-70%) during the first set of national non-pharmaceutical interventions including school closures in late March (see Materials and Methods, Fig 1C). Travel had an overall rate of decay of 13% during this first wave of COVID-19 cases and NPIs from March-May 2020. These reductions were largely driven by decreases in inter-province travel with the majority (44/51; one route did not have recorded trips) of routes showing decreases, but over a range of values (6-96%) (Supplement Fig 1). Interesting, travel between Copperbelt and Southern provinces, a moderately trafficked route, saw an increase in 45% of daily trips during this period, despite overall decreases nationally. The top three most traveled provincial routes saw some of the largest reductions (Central – Lusaka: 69%, Lusaka – Southern: 78%, Central – Copperbelt: 89.9%) which corresponds to a decrease up to 2,208 daily trips per route (range 802-2,208).

Less traveled routes overall experienced a lower relative reduction in travel (on average 30%, see Supplement Fig 1) with a few (7/51) locations also showing an increase in travel. Inter-district travel showed similar patterns to inter-province travel with reductions of 1-98% (IQR: 37-73%) relative to baseline. Similarly, the most frequently traveled district routes during baseline were also those that saw the greatest relative reductions in travel (average 70%, IQR: 68-75%) (Fig 1C).

Travel within provinces also saw reductions, but to a lesser degree than inter-province (Fig 2A-B). Overall, intra-province travel decreased during the first wave by 23-52% of baseline (Fig 2B). The provinces where the largest number of trips were recorded saw fewer reductions in travel after restrictions were implemented. For example, travel within Lusaka, Copperbelt, and Southern Provinces saw reductions of 36%, 32%, and 23% during Wave 1, respectively. In contrast, intra-province travel within provinces with the fewest number of trips recorded saw larger reductions than those in more traveled routes (North-Western and Muchinga, experienced reductions of 42% and 47%, respectively). Intra-district mobility did not see the same reductions as inter-district mobility (reductions of 3-80% (IQR: 24-46%) during Wave 1 compared to baseline (only two intra-district routes saw increases). This is further illustrated by the ratio of inter- to intra-province travel which fell 64% during the first wave (see Fig 2C), but with differences across routes. During Wave 1, more frequently traveled routes had *greater* reductions in *inter*-province travel, whereas more frequented routes had *reduced* reductions in *intra*-province travel. This is likely due to NPIs such as stay-at-home orders, restrictions on public events, and limitations on gatherings put into place at the beginning of the first wave, discouraging inter-province travel.

**Fig 2.**
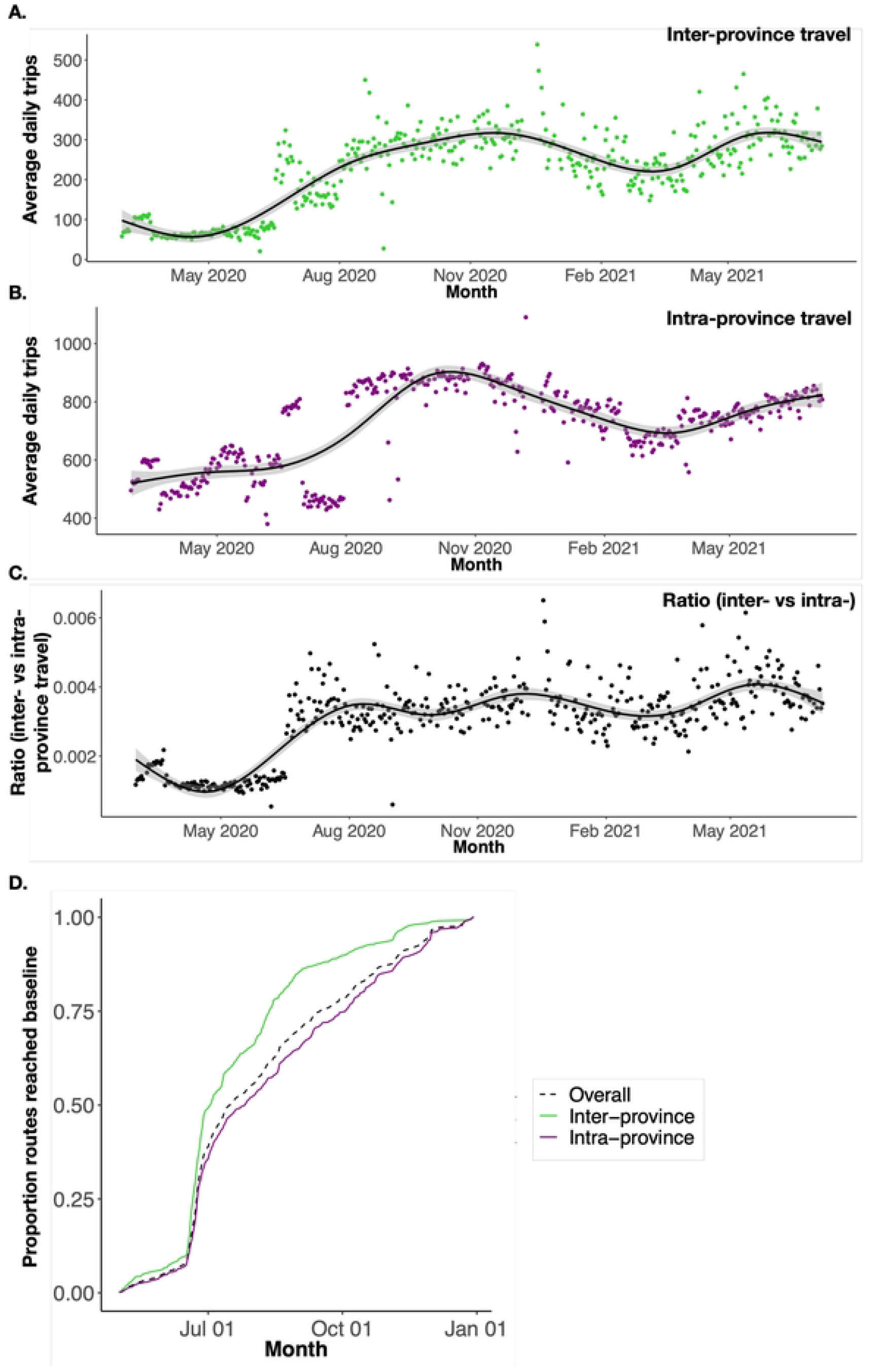
Inter-province versus intra-province daily trips and return to baseline mobility levels following implementation of NPIs. **(A)** The average daily trips between and **(B)** within provinces from March 2020 to July 2021. Points represent daily average mobility estimates and solid line is the smoothed conditional average of the points with the 95% confidence interval shaded. **(C)** The ratio of inter- to intra-province travel calculated as the number of trips between provinces divided by the number of trips within a province. Points represent the daily ratio of inter-versus intra-province travel and the solid line represents the smoothed conditional average of the points. **(D)** The proportion of inter- and intra-province routes that have recovered to baseline values over time (in months). Baseline mobility and recovery time was calculated for induvial routes. Overall (dashed line) represents the average across inter- and intra-province routes

### Mobility returned to pre-pandemic levels within a few months

Following the lift of the strictest NPIs on 08 May 2020, the number of trips and routes of travel returned to baseline by July 2020. Intra-province travel recovered faster than inter-province travel (intra: average = 2.5 months, IQR: 1.5 – 3.2 months; inter: average = 3.3, IQR 1.6 – 4.9 months) (Fig 2D). The top 10 most traveled routes recovered faster (average 2 months) whereas the 10 less frequented routes recovered slower (average 2.43 months); however, differences in recovery between most and least commonly traveled routes was not significant (p-value = 0.29). Across all routes, 15% of district routes took 6 months or greater to return to baseline. However, these routes were rarely traveled during the baseline period with an average of 1.97 trips per day and were likely not heavily trafficked during the pandemic. Once mobility returned to baseline following the lift of NPIs during the first wave, mobility remained stable during subsequent waves and large reductions in inter- and intra-province mobility was not observed.

### Mobility did not decrease in response to COVID-19 case reports

Unlike other settings globally, COVID-19 cases were less common in Zambia with few districts (25/116) reporting at least one confirmed case by the end of May 2020. Among these districts, including some with the first and highest number of cases reported (Lusaka, Lusaka Province and Nakonde, Muchinga Province) we did not see any differences in the reduction in mobility or return to baseline travel compared nationally (p-value = 0.86). Further, during the subsequent waves starting in July 2020 and Jan 2021, we did not see any additional reductions in mobility suggesting COVID-19 case counts did not impact differences in mobility (Fig 3A).

**Fig 3.**
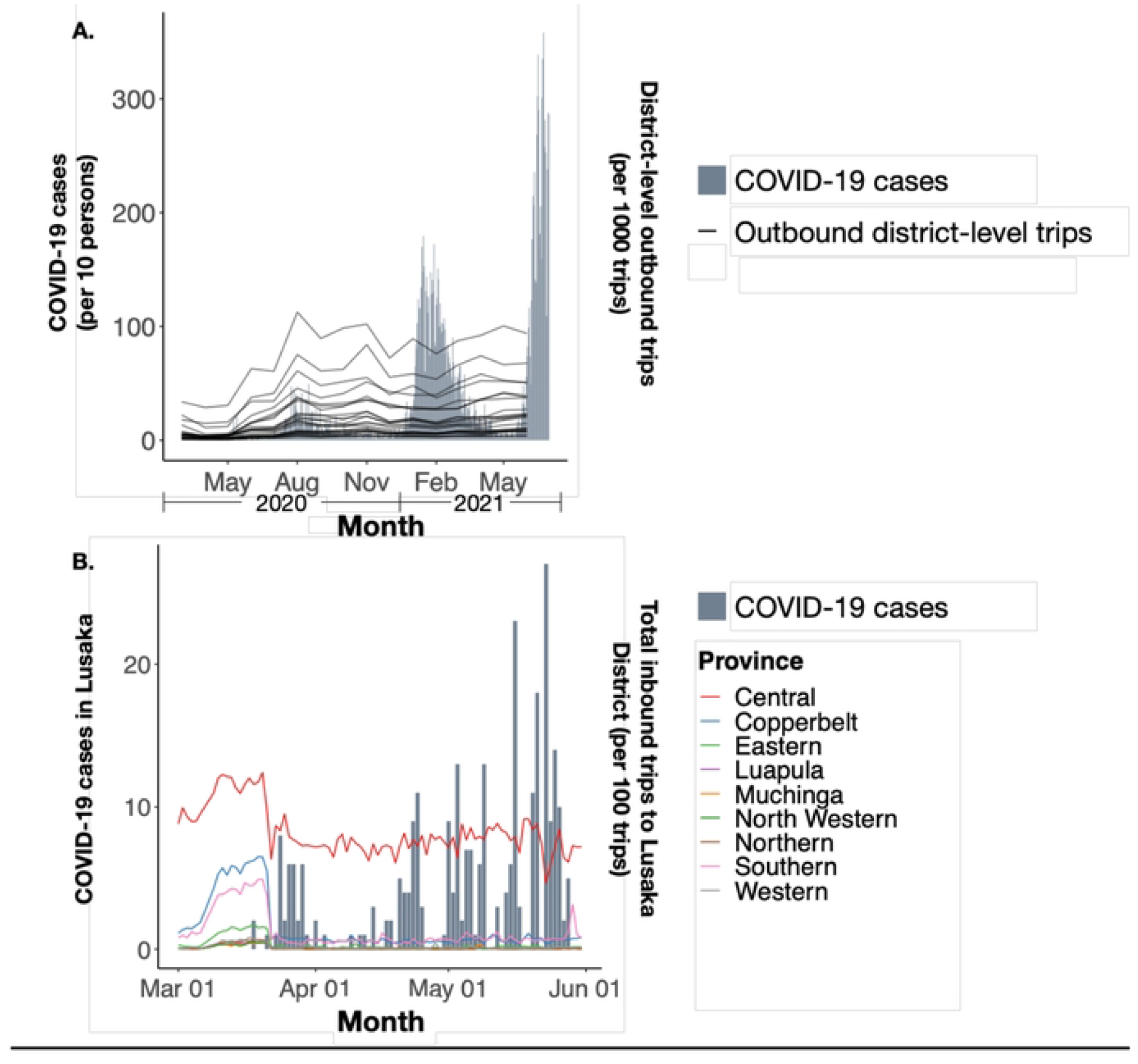
Mobility patterns during waves of COVID-19 cases at national and sub-national levels. **(A)** Total district-level outbound trips among the 24 districts who reported having at least one COVID-19 case during Wave 1. Grey bars represent national COVID-19 cases from March 2020-June 2021. **(B)** Total daily inbound trips to Lusaka District from other Provinces (excluding Lusaka Province) from 01 March 2020 – 01 June 2021. Grey bars represent daily COVID-19 cases in Lusaka district during Wave 1 (01 March 2020 – 31 May 2020).

Given variability in testing and reporting across Zambia at the start of the pandemic, we focused our subsequent analyses in Lusaka where testing was more readily available. During the period following the first case report in Lusaka on 18 March 2020 and the end of Wave 1, traffic from other provinces into Lusaka decreased, on average, 19.6% compared to baseline (Fig 3B); however, reductions into Lusaka did not significantly differ from travel in other districts and provinces (p-value = 0.08). Travel to Lusaka District other districts within Lusaka Province decreased by 67% on average compared to baseline; directly following the first case report, mobility dropped across all districts within Lusaka Province and remained low during the first wave until they eventually rebounded around May and June 2020 where it remained stable for the duration of the year (Fig 3B). Further, intra-province travel in Lusaka Province had significantly greater reductions in district-level mobility compared to other provinces (p = 0.001).

### Mobile phone data better illustrates district-level mobility compared to Google Mobility Reports

Google Mobility Reports have served as a popular source of data for analyses aiming to incorporate human mobility data given their global availability. However, how these data compare to mobile phone data has not been widely explored in SSA. Overall, Google Mobility Reports estimate lower magnitudes of change in mobility compared to the mobile phone data used in this analysis, however, a similar trend is consistent across datasets during Wave 1 (Fig 4A). It is important to note that Google Mobility Reports are available nationally, and thus cannot be used to understand district or province level heterogeneity. Additionally, the mobile phone data report larger decreases in less frequently traveled routes, which is likely driving national reductions to higher magnitudes. Since Google Mobility Reports are national, the impacts of less traveled, subnational routes may not be contributing to overall reductions as dramatically as seen with the mobile phone data. Lastly, most Google application users are likely in more urban areas, which, despite having great reductions during Wave 1, still maintained a higher level of mobility; this prevents the data from accounting for less frequently traveled trips to rural areas.

**Fig 4.**
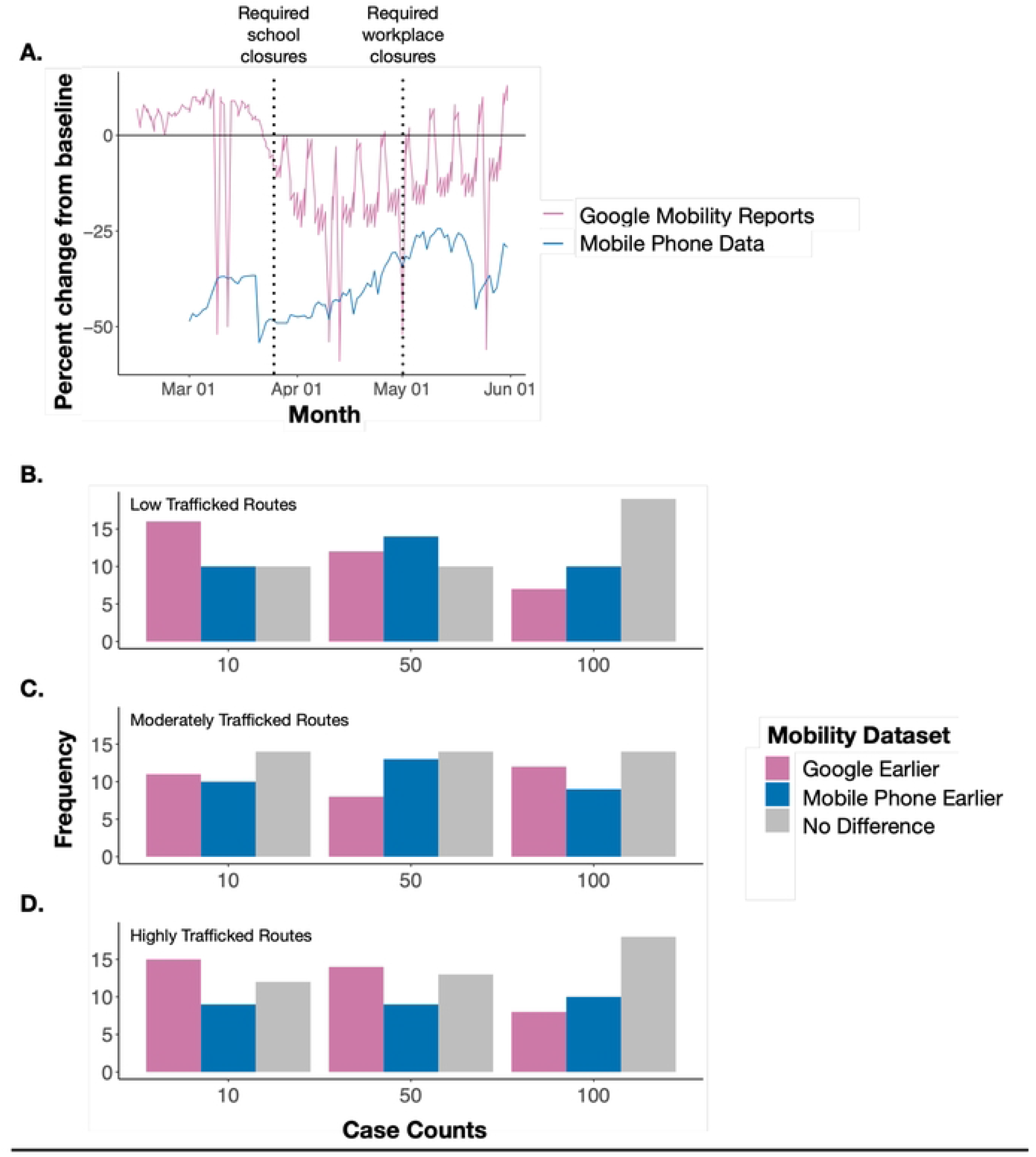
Comparing Mobile Phone Data and Google Mobility Reports in overall mobility reporting and COVID-19 case predictions. **(A)** The percent change from baseline mobility during Wave 1 (March – May 2020) for Google Mobility reports (Pink) and Mobile Phone Data (blue). Mobile Phone Data include all intra-province trips. Dashed lines represent implementation of key NPIs such as school and workplace closures. Large dips shown by Google Mobility reports are national holidays where there were large reductions in mobility to and from workplaces. The frequency of districts in which either Google or Mobile Phone data predicted a faster rise in SARS-CoV-2 cases across **(B)** low (≤2,250 total baseline trips), **(C)** moderately (2,250-7,630 total baseline trips), and **(D)** highly trafficked routes (≥7,630 total baseline trips).

To further illustrate the impact of using both types of data, we simulated transmission dynamics of SARS-CoV-2 during 2020 using mobility data from Google or mobile phone data. We performed a simple simulation reflective of the overall patterns of the initial stages of the pandemic in Zambia where initially only cases were reported in Lusaka district in March 2020. Further, we investigated the time till at least 10 cases were imported to all other districts either informed by Google or mobile phone data. Despite differences in the reductions in travel between the data sets, there were no large differences in time to the first 10 cases using Google data or mobile phone data (Fig 4B-D, Supplement Fig 3A-B). Across all districts, Google data projected earlier case counts in 39% of districts, mobile phone data projecting earlier case counts in 27%, and 24% of districts had no difference between datasets used. Despite a small difference, these results align with our expectations that Google would produce earlier estimates of cases rising given that the Google data suggested smaller reductions in mobility, which would have contributed to an increased risk for introduction events in other districts. We further stratified by travel frequency to each district defined as low, moderate, or highly trafficked districts (Low: ≤2,250 total baseline trips, Moderate: 2,250-7,630 total baseline trips, and High: ≥7,630 total baseline trips). We see that as the case incidence increases, the difference in time to cases becomes less prevalent between datasets (Fig 4B-D).

## Discussion

Human mobility has been widely impacted from global to sub-national scales throughout the course of the pandemic. Dissecting the patterns of human mobility during the pandemic can help quantify the impact of NPIs on mobility as well as how changes in travel can influence SARS-CoV-2 transmission. Here, we aimed to evaluate human mobility patterns in Zambia, which to date, has largely been unexplored. Overall, mobility decreased during the first Wave (March – May 2020) following the implementation of strict NPIs including school and workplace closures, stay at home orders, and restrictions on gatherings. During this time, COVID-19 cases were rising, but were maintained at lower levels compared to other countries in Africa. [18] However, following the first wave, mobility returned to baseline and remained at baseline levels for the duration of 2020. Despite subsequent waves of COVID-19 and the continuation of less stringent NPIs, mobility did not drop at the same magnitudes seen during Wave 1. Further, COVID-19 cases continued to rise and fall while mobility remained at baseline levels suggesting increases in cases did not produce reductions in mobility, rather enforced and stringent NPIs played a larger role in influencing mobility, especially during Wave 1.

Estimating human mobility during the pandemic has been of large interest and companies such as Google and mobile service providers have been important sources of data. In this analysis, we highlight the differences between these two sources and conclude that mobile phone data contributes more spatially refined data by collecting data based off phone towers and not app usage, which is beneficial for mapping mobility in more rural areas. Conversely, Google Mobility Reports rely on app usage to illustrate weekly national peaks and dips, categorized by type of mobility, that is not observed in the mobile phone data. Overall reductions suggested by Google Mobility Reports likely underestimate reductions in less frequently traveled routes as well as reductions seen nationally. The accessibility of these data makes it an attractive source for public health officials. However, these data may be biased; mobile phone data relies on device ownership and Google data is reliant upon device owners having their location services on and utilization of Google Maps. Overall, both types of data have benefits and drawbacks, but both sources have been critical to aid researchers and public health officials in mapping the impact of NPIs and simulating trajectories of SARS-CoV-2 transmission.

There are a few limitations of this analysis. First, the data comes from only one of three operational mobile phone providers in Zambia and, though having nation-wide coverage, we do not know the proportion of subscribers in country and may not be representative of the entire population. Second, we did not have data available for 2019, or pre-pandemic, time periods. In this analysis we used a period where mobility leveled out (September – December 2020) to serve as a proxy for baseline mobility, which may be an underestimation of true baseline mobility values. Lastly, due to Google Mobility Reports only providing national estimates, we were unable to extract actual values for district routes; instead, we took a proportion of the baseline data (September – December 2020) based on the national reduction estimates.

Overall, strict NPIs impacted mobility in the early stages of the pandemic. Given the risk of variants, it is important to continue the consideration of NPIs to curb COVID-19 outbreaks. Human mobility plays a large role in the propagation of infectious diseases and continues to be a key element in strategies aimed at reducing transmission. While SARS-CoV-2 transmission is likely to transition into an endemic disease, it remains critical to consider the intersection of NPIs and human mobility as we navigate through the future of the COVID-19.

## Data Availability

Data is available by request to the authors (included as a URL for review).

## Supporting Information

**Supp Fig 1. Total daily trips between provinces recorded from March 2020 to July 2021**. Each row represents the starting province and each line represents the total number of daily trips to a destination province. Intra-province trips (trips occurring within the same province) are excluding from this figure.

**Supp Fig 2. Maps displaying the logged values of trip counts for the top 10% most frequently traveled routes stratified by wave**. Values were normalized by scaling to minimum and maximum values. **(A)** Top 10% of trips during Wave 2 (June-August 2020) and **(B)** Top 10% of trips during Wave 3 (January-March 2021).

**Supp Fig 3. Time series of predicted COVID-19 cases by district for Google or Mobile Phone data. (A)** Time series of infected persons using Google versus Mobile Phone data with Lusaka District (blue-green line) included. **(B)** Time series of infected persons using Google versus Mobile Phone data <u>excluding</u> Lusaka District.

## References

[1] Hale T, Angrist N, Goldszmidt R, Kira B, Petherick A, Phillips T, et al. A global panel database of pandemic policies (Oxford COVID-19 Government Response Tracker). Nat. Hum. Behav. 2021;5(4):529–38.

[2] Brauner JM, Mindermann S, Sharma M, Johnston D, Salvatier J, Gavenčiak T, et al. Inferring the effectiveness of government interventions against COVID-19. Science 2021;371(6531).

[3] Hu T, Wang S, She B, Zhang M, Huang X, Cui Y, et al. Human mobility data in the COVID-19 pandemic: characteristics, applications, and challenges. Int. J. Digit. Earth 2021;14(9):1126–47.

[4] Ilin C, Annan-Phan S, Tai XH, Mehra S, Hsiang S, Blumenstock JE. Public mobility data enables COVID-19 forecasting and management at local and global scales. Sci. Rep. 2021;11(1):13531.

[5] Mulenga LB, Hines J, Fwoloshi S, Chirwa L, Siwingwa M, Yingst S, et al. Prevalence of SARS-CoV-2 in six districts in Zambia in July, 2020: a cross-sectional cluster sample survey. Lancet Glob. Heal. 2021;9(6):e773–81.

[6] Ogunleye O, Basu D, Mueller D, Sneddon J, Seaton RA, Yinka-Ogunleye, AF, et al. Response to the Novel Corona Virus (COVID-19) Pandemic Across Africa: Successes, Challenges, and Implications for the Future. Frontiers in Pharmacology 2020;11(11):1205.

[7] Kishore K, Jaswal V, Verma M, Koushal V. Exploring the utility of google mobility data during the COVID-19 pandemic in India: Digital epidemiological analysis. JMIR Public Heal. Surveill. 2021;7(8):e29957.

[8] Africa CDC. Africa CDC COVID-19 Vaccine Dashboard [Internet]. Updated 2022; Accessed 13 Apr 2022. Available from: https://africacdc.org/covid-19-vaccination/

[9] Our World in Data. COVID-19 Data Explorer - Our World in Data [Internet]. Updated 2022; Accessed 13 Apr 2022. Available from: https://ourworldindata.org/covid-vaccinations

[10] WHO Africa. Key lessons from Africa’s COVID-19 vaccine rollout [Internet]. Updated 2021; Accessed 20 Jan 2022. Available from: https://www.afro.who.int/news/key-lessons-africas-covid-19-vaccine-rollout

[11] Bushman M, Kahn R, Taylor BP, Lipsitch M, Hanage WP. Population impact of SARS-CoV-2 variants with enhanced transmissibility and/or partial immune escape. Cell 2021;184(26):6229–42.

[12] oogle. Google COVID-19 Community Mobility Reports [Internet]. 2020; Accessed 25 June 2021. Available from: https://www.google.com/covid19/mobility/

[13] Dong E, Du H, Gardner L. An interactive web-based dashboard to track COVID-19 in real time. The Lancet Infectious Diseases 2020;20(5):533–4.

[14] Kristensen K, Bell B, Skaug H, Magnusson A, Berg C, Nielsen A. TMB: Template Model Builder: A General Random Effect Tool Inspired by ‘ADMB’ [Internet]. Updated 2022; Accessed 14 Apr 2022. Available from: https://cran.r-project.org/package=TMB

[15] Finkenstädt BF and Grenfell BT. Time series modelling of childhood diseases: A dynamical systems approach. J. R. Stat. Soc. Ser. C Appl. Stat. 2000;49(2):187–205.

[16] Iyaniwura SA, Rabiu M, David JF, Kong JD. The basic reproduction number of COVID-19 across Africa. PLoS One 2022;17(2):e0264455.

[17] Khan ZS, van Bussel F, Hussain F. A predictive model for COVID-19 spread – with application to eight US states and how to end the pandemic. Epidemiol. Infect. 2020;148:E249.

[18] Africa CDC. Novel Coronavirus (2019-nCoV) Global Epidemic – 31 March 2020. Africa CDC COVID-19 Brief 2020;11:1–7.

